# Antibody response to *Plasmodium vivax* in the context of Epstein-Barr virus (EBV) co-infection: A 14-year follow-up study in the Amazon rainforest

**DOI:** 10.1101/2024.09.25.24314386

**Authors:** Luiz F. F. Guimarães, Bárbara A. Rodrigues, Michelle H. F. Dias, Matheus G. Barcelos, Maria F. A. Nascimento, Sâmick L. Moreira-Nascimento, Sofia L. Afonso, Barbara G. S. Abreu, Jaap. M. Middeldorp, Francis B. Ntumngia, John H. Adams, Camila Fabbri, Stefanie Lopes, Cor J. F. Fernandes, Flora S. Kano, Luzia H. Carvalho

## Abstract

**Background:** To develop an effective vaccine against *Plasmodium vivax*, the most widely dispersed human malaria parasite, it is critical to understand how coinfections with other pathogens could impact malaria-specific immune response. A recent conceptual study proposed that Epstein-Barr virus (EBV), a highly prevalent human herpesvirus that establishes lifelong persistent infection, may influence *P. vivax* antibody responses. Here, it was investigated whether EBV could impact the longevity of humoral immune response to *P. vivax*.

**Methodology/principal findings:** A 14-year follow-up study was carried out among long-term *P. vivax*-exposed Amazonian individuals (272, median age 35 years), and included 9 cross-sectional surveys at periods of high and low malaria transmission. The experimental approach focused on monitoring antibodies to the major blood-stage *P. vivax* vaccine candidate, the Duffy binding protein region II (DBPII-Sal1), including a novel engineered DBPII-based vaccine targeting conserved epitopes (DEKnull-2). In parallel, the status of EBV infection was determined over time by the detection of circulating EBV DNA (EBV-DNAemia) and EBV-specific antibodies to lytic (VCAp18) or latent (EBNA1) antigens. Regardless of the malaria transmission period, the results demonstrated that one or multiple episodes of EBV-DNAemia did not influence the longevity of DBPII immune responses to both strain-specific (Sal-1) or strain-transcending (DEKnull-2) antibodies. Also, the average time in which DBPII-responders lost their antibodies was unrelated to the EBV serostatus. Considering all malaria cases detected during the study, there was a predominance of *P. vivax* mono-infection (76%), with a positive association between malaria infection and detectable EBV-DNAemia.

**Conclusions/significance:** In an immunocompetent *P. vivax*-exposed adult population neither sporadic episodes of EBV-DNAemia nor antibody responses to lytic/latent EBV antigens influence the longevity of both strain-specific and strain-transcending DBPII immune responses. Further studies should investigate the role of acute *P. vivax* infection in the activation of EBV replication cycle.

## Introduction

Despite the decline in the global burden of malaria, the progress towards disease control has stagnated in recent years, underscoring the fragility of hard-won gains against malaria [1]. In the case of *Plasmodium vivax*, the most widespread human malaria parasite, plateauing gains and areas with an increased burden highlight the fact that the challenges on the road to malaria elimination are greater than expected [2]. Key discoveries in *P. vivax* biology, including an endosplenic life cycle associated with recurrent chronic infections, pose additional challenges to control and elimination of the parasite [3, 4]. Concurring, the proportion of malaria caused by *P. vivax* has been increasing in co-endemic regions where intensive malaria-control activities have successfully reduced the incidence of *P. falciparum* [5].

Although under investigated, *P. vivax* in a scenario of co-infection with viruses commonly circulating in endemic areas can worsen the impact of these diseases on public health, reducing the effectiveness of therapeutic and prophylactic measures [6–11]. With continuing interest in the elimination of *P. vivax* infections, and to the development of an effective vaccine against this parasite, it is necessary to understand how viral infections could impact the immune response induced by potential vaccine candidates. Particularly, because there are only a few *P. vivax* vaccines in clinical trials [12].

In experimental models, it has been proposed that gamma herpesvirus co-infection with malaria may suppress anti-parasitic humoral immunity [13]. Given that the B-cell compartment is the primary niche for Epstein-Barr virus (EBV), a highly prevalent human gamma herpes virus [14], it is plausible to speculate that the humoral immune response to malaria may be altered during EBV co-infection [13]. In a conceptual case-control study, we provided evidence that antibody levels against major *P. vivax* vaccine candidates were generally lower in malaria-exposed individuals whose viral DNA was persistently detected in the peripheral blood [15]. To further investigate whether EBV could impact the long-term humoral immune response to *P. vivax*, we took advantage of a 14-year follow-up study among individuals exposed to malaria in the Amazon rainforest, focusing on a major blood-stage *P. vivax* vaccine candidate, Duffy binding protein region II (DBPII).The experimental approach included retrospectively analyzing the relationship between the long-term antibody response to DBPII-related antigens in the presence (or absence) of a sustained response to EBV, as determined by the detection of circulating EBV DNA (EBV-DNAemia) or EBV-specific antibody responses to the viral capsid antigen P18 (VCAp18) or EBV nuclear antigen 1 (EBNA1).

## Material and methods

### Area and study population

The study was carried out in the agricultural settlement of Rio Pardo (1°46’S—1°54’S, 60° 22’W—60°10’W), Presidente Figueiredo municipality, Northeast of Amazonas State in the Brazilian Amazon region (Fig 1A). The study site and malaria transmission patterns were described in detail elsewhere [16–18]. In this area, malaria transmission is considered hypo to mesoendemic, and most residents were natives of the Amazon region. Inhabitants of the settlement live on subsistence farming and fishing along the small streams. In the study area, *P. vivax* transmission is predominant over *P. falciparum* (Fig 1B) and, since 2010, *P. vivax* has been responsible for all clinical cases of malaria. Malaria transmission decreased drastically in recent years, and as of 2020, no new clinical cases have been reported (Fig 1B).

**Fig 1.** Study site and methodological approach according to the temporal distribution of malaria cases during the 14-year follow up period (2008-2022). A) The study site, Rio Pardo settlement (dark green), in the Amazonas State (AM) of the Brazilian Amazon region (malaria-endemic area, light green). B) Monthly-time series of malaria cases in the study area, including periods of high (I and III, light grey) and low (II and IV, dark grey) malaria transmission. Malaria cases (microscopy-positives) were reported by the Brazilian Epidemiological Surveillance System for Malaria (SIVEP-Malaria), with *P. falciparum* (red) and *P. vivax* (blue) cases plotted per month. C) Study design included nine cross-sectional surveys which were carried-out between 2008 and 2022. Number of participants enrolled per cross-sectional survey is shown inside the circles above (cross-sectional, n), and number of eligible participants for the current study is inside the circles below (eligible participants, n).

### Study design and cross-sectional surveys

A population-based open cohort study was initiated in November 2008 and included three cross-sectional surveys carried-out at six-month intervals (baseline, 6 and 12 months) as previously reported [15, 16]. Briefly, (i) interviews were conducted through a structured questionnaire to obtain demographical, epidemiological, and clinical data; (ii) physical examination, including body temperature and spleen/liver size were recorded according to standard clinical protocols; (iii) venous blood was collected from individuals aged five years or older (EDTA, 5 mL); and (iv) examination of Giemsa-stained thick blood smears for the presence of malaria parasites by light microscopy. The geographical location of each dwelling was recorded using a hand-held 12-channel global positioning system (GPS) (Garmin 12XL, Olathe, KS, USA) with a positional accuracy of within 15m. Additional cross-sectional surveys were conducted during the 6th to 14th years of the study, specifically on the following dates: October 6–13, 2014 (6th year), July 6–11, 2015 (7th year), July 18–27, 2017 (9th year), August 4–16, 2019 (11th year), November 28–December 10, 2021 (13th year), and June 6–16, 2022 (14th year). During the long-term follow up study, the number of malaria cases fluctuated in the study area, reflecting period of high (I and III) and low (II and IV) malaria transmission (Fig 1B). The current study involved a retrospective approach, and the methodological strategy was designed to include samples from all nine cross-sectional surveys without any new recruitment involved. For this, a database (EpiData software; http://www.epidata.dk) contained all individual information was accessed between June 27 of 2022 until July 30, 2024; in the database individual data were not linked to participants’ identity as participants were assessed by codes. The non-eligible criteria were (i) refusal to sign the informed consent; (ii) young children (<5 years); (iii) pregnant women; (iv) any other morbidity that could be traced; (iv) individuals who were unable to be recruited during at least two cross-sectional surveys, (iv) individuals whose biological samples (DNA and plasma) were unavailable. A total of 272 individuals were eligible to the current study (Table 1), and they matched the original population for age, sex, malaria exposure [17]. The number of eligible participants selected per cross-sectional surveys was included in the Fig 1C.

**Table 1.** Demographic and epidemiological data of participants at the time of enrollment.

| Variable | At enrollment<br>(n=272) |
| --- | --- |
| Age, years - Median (IQR<sup>1</sup>) | 35 (14-51) |
| Gender - male: female | 1.12:1 |
| Riverine population, n (%) | 143 (53%) |
| Years of malaria exposure - Median (IQR<sup>1</sup>) | 29 (15-48) |
| Years of residence in Rio Pardo - Median (IQR<sup>1</sup>) | 19 (12-21) |

<sup>1</sup>IQR, the Interquartile range

The ethical and methodological aspects of this study were approved by the Ethical Committee of Research on Human Beings from the René Rachou Institute (CAAE: 96098618.9.0000.5091), according to the Resolutions of the Brazilian Council on Health (CNS-196/96 and CNS-466/2012). Formal written consent was obtained from all participants, either from the participants themselves, their next of kin, caregivers, or guardians on behalf of child participants. Data are separated from personal identifiers through use of a code.

The current study was conducted according to Laboratory biosafety and biosecurity policy guidelines of the Oswaldo Cruz Foundation (FIOCRUZ, Ministry of Health, Brazil (http://www.fiocruz.br/biosseguranca/Bis/manuais/biosseg_manuais.html).

### Laboratory diagnosis of malaria

At the time of blood collection, all individuals were submitted to a finger-prick for malaria diagnosis by light microscopy. The Giemsa-stained thick blood smears were prepared and examined by experienced local microscopists, according to the malaria diagnosis guidelines of the Brazilian Ministry of Health. Species-specific real-time PCR assay was conducted essentially as previously described [19]. For this, genomic DNA was extracted from either whole blood samples collected in EDTA, or from dried blood spots on filter paper using the Puregene blood core kit B (Qiagen, Minneapolis, MN, USA) or the QIAmp DNA mini kit (Qiagen), respectively, according to manufacturers’ instructions.

### Recombinant blood stage *P. vivax* proteins and IgG antibodies detection

#### DBPII-related antigens

DBPII-Sal1, a recombinant Duffy binding protein region II (DBPII) [20] including amino acids 243–573 of the Sal1 reference strain, and recombinant DEKnull-2 [21], an engineered DBPII immunogen, were expressed as a 39kDa 6xHis fusion protein and properly refolded, as previously described [21, 22]. Conventional Enzyme-Linked Immunoassays (ELISA) for *P. vivax* IgG antibodies were carried out using *P. vivax* blood-stage recombinant proteins as previously described [23], with serum samples at a dilution of 1:100. Recombinant proteins were used at a final concentration of 3 μg/mL. The results were expressed as ELISA reactivity index (RI), calculated as the ratio of the mean optical density (OD at 492 nm) of each sample to the mean OD plus three standard deviations of negative control plasma samples from 30 individuals living in a nonendemic area of malaria (Belo Horizonte, Minas Gerais, Brazil) and who have never been exposed to malaria transmission (unexposed volunteers). Values of RI > 1.0 were considered seropositive.

### EBV DNA detection by real-time PCR

The PCR primers for this assay were previously selected in the single-copy BALF-5 gene encoding the viral DNA polymerase [24]; the upstream and downstream primer sequences were 50-CGGAAGCCCTCTGGACTTC-30 and 50-CCCTGTTTATCCGATGGAATG-30, respectively, with a fluorogenic probe (50-TGTACACGCACGAGAAATGCGCC-30) with a sequence located between the PCR primers. Detectable DNA from EBV was identified by a real-time PCR assay as previously described [15]. Briefly, DNA from whole blood samples collected in EDTA was extracted using Puregene blood core kit B (Qiagen, Minneapolis, MN, USA). The PCR reaction was performed using a mixture containing 1μL of DNA, 0.2 μM each primer, 0.1 μM fluorogenic probe, and 5 μL of TaqMan Master Mix (PE Applied Biosystems), and the PCR cycle was performed as follows: 2 min at 50°C, 10 min at 95°C, and 40 cycles of 15 s at 95°C and 1 min at 60°C. The TaqMan Master mix (PE Applied Biosystems) was used for all reactions. For all PCR analyses, water was used as negative control, and DNA samples from individuals with history of mononucleosis were used as positive control. Samples were defined as negative if the CT values exceeded 40 cycles.

### EBV antigens and serostatus

EBV-specific antibodies were detected using two synthetic peptides covering immunodominant epitopes of the viral capsid antigen P18 (VCAp18 [BFRF3]) and EBV nuclear antigen 1 (EBNA1 [BKRF1]) [25, 26]. Both synthetic peptides were provided by Dr. J. M. Middeldorp (VU University Medical Center, Amsterdam, Netherland). For the assessment of the levels of antibodies against lytic (VCAp18) and latent (EBNA1) EBV antigens, we used synthetic peptide-based ELISA assays as previously described [15]. Briefly, each peptide was used at final concentration of 1μg/mL with plasma samples diluted 1:100. IgG reactivities were determined using commercial anti-human IgG secondary antibody conjugated to horseradish peroxidase (HRP) (Sigma-Aldrich). For each experiment, plasma samples from subjects with or without a history of mononucleosis infection who had previously been screened for the presence or absence of EBV-specific antibodies were included as positive (EBV seropositive) or negative controls (seronegative), respectively. All OD 450nm values were normalized by subtracting the value for 1:100-diluted EBV negative sera used in duplicate in each ELISA run. ELISA’s cutoffs were previously established as [15]: (i) 0.38 for VCA IgG (82% sensitivity; 83% specificity), (ii) 0.20 for EBNA1 IgG (75% sensitivity; 100% specificity).

### Statistical analysis

A database was created using EpiData software (http://www.epidata.dk). The graphics and the statistical analyses were performed using GraphPad Prism version 9.5.0 - GraphPad Software (La Jolla, California, USA) and Stata software version 12.0 - Stata Corp (College Station, Texas, USA).

The Shapiro-Wilk test was performed to evaluate normality of variables. For the statistical analyses, the antibody response was defined either as a continuous variable (the levels of antibody response) or as a binary categorical variable (proportion of seropositive individuals); differences between medians were tested by Mann-Whitney test or Kruskal-Wallis as appropriate; differences in proportions were evaluated by chi-square test or Fisher’s exact test, as appropriated. Spearman’s correlation matrices were used to nonparametric measure of rank correlation. For statistical purposes, the amount of time participants contributed to this open cohort study was estimated in persons-month, with the sum of total time contributed by all subjects (months) used to calculate the incidence densities of the events of interest. Kaplan-Meier curves of survival were used to estimate the probability of remaining seropositive for *P. vivax* as function of the profile of EBV infection (serostatus and EBV-DNAemia). For that, the average duration of seropositivity over the 14th-years period was defined when 50% of responders lost their antibodies. Logistical regression models were constructed to investigate *P. vivax* antibodies clearance (rates adjusted per 100 persons-month) according to the profile of EBV infections, with the relative risk (95% CI) of losing *P. vivax* antibody response estimated according to the first seronegative episode. For all analyses, an alpha error of 0.05 was considered.

## Results

### *Plasmodium vivax* infections and circulating EBV DNA during the follow-up

In this long-term follow-up study, malaria blood-stage infections varied significantly, but with a predominance of sub-patent (PCR-positive) over patent (microscopy-positive) infections (Fig 2A). All malaria infections detected by microscopy were identified as *P. vivax* mono-infections, with all but one (11 of 12) detected during the first year of the follow-up (Period I). Submicroscopic malaria infections were also predominant in the first year of follow-up, with frequency ranging from 9 to 22% (Fig 2A). In subsequent cross-sectional surveys, lower frequencies of PCR-positive samples were detected (3-4% and 4-6% during periods II and III, respectively), with no cases of malaria detected at the end of the study (IV) (Fig 2A). Considering all malaria cases detected during the study (n=113), there was a predominance of *P. vivax* mono-infection, accounting for 76% (n=86) of the total malaria infections, compared to 12% (n=14) of *P. falciparum*, and 12% (n=13) of mixed infections by these two *Plasmodium* species (S1 Fig).

**Fig 2.** Malaria infections and EBV-DNAemia during the follow-up-period. A) Frequencies of malaria positive samples detected by microscopy (red bars) or species-specific PCR-based protocol (light pink bars) and B) Frequencies of circulating EBV DNA (EBV-DNAemia) detected by virus-specific PCR-based protocol (lilac bars). Each bar represents a cross-sectional survey, with dashed lines representing the periods of high (I and III) and low (II and IV) malaria transmission, as described in the legend of Fig 1B. C) A comparison between the proportion of malaria infections and EBV-DNAemia over time. D) Representation of a Spearman correlation coefficient, in which each dot represents the intersection between the frequency of malaria infections and detectable EBV-DNAemia by cross-sectional survey. Numbers at the bottom of each bar (A, B) represent the number of participants per survey (n survey) and the total number of participants per period (n period).

On the first period of follow-up, the proportion of individuals who had circulating viral DNA (EBV-DNAemia) ranged from 28 to 37% (Fig 2B). After that, EBV-DNAemia episodes decreased substantially (Fig 1C), except for the beginning of period III (9-years of the follow-up), a time of high malaria transmission in the study area (Fig 1B). It is noteworthy that the lowest frequencies of EBV DNA positivity were found during the II (11 to 14%) and IV (2 to 3%) periods of the study (Fig 2B), corresponding to periods of lowest malaria transmission in the area (Fig 1B). In the study population, episodes of EBV-DNAemia were transiently detected in most individuals, which excluded the possibility of categorizing individuals as having long-term persistent EBV-DNAemia. Only seven out of 272 individuals had long-term EBV-DNAemia (positive samples from baseline to at least the II period); four of them (343, 272, 179, 276) had malaria infections at some point in the study (S1 Fig). During the follow-up, a strong positive association was detected between malaria infections and EBV-DNAemia (r = 0.7269; p = 0.0035) (Fig 2D). Similar results were found when only *P. vivax* monoinfections were analyzed (r = 0.6522; p = 0.0085, data not shown).

### Episodes of EBV-DNAemia and longevity of naturally acquired antibodies against *Plasmodium vivax*

In the study population, the profile of antibody response to DBPII-based antigens decreased over time, with a more prolonged response to DEKnull-2 in period II (low transmission) as compared with DBPII (Sal1) (S2 Fig); at the same time, the levels of anti-DBPII antibodies were lower than anti-DEKnull-2. To further evaluate whether episodes of EBV-DNAemia (none, one, or multiple) could influence the persistence of strain-specific (Sal1) or strain-transcending (DEKnull-2) DBPII antibody repertoire, Kaplan-Meier’s survival curves were constructed. In general, the results demonstrated that the persistence of antibody responses to either DBPII-Sal1 or DEKnull-2 was similar between individuals who had none, one or multiple episodes of EBV-DNAemia (Fig 3). More specifically, considering the subgroups of DBPII-Sal1 responders, the average time in which 50% of responders lost their antibody response ranges from 156 to 168 months (S1 Table). The average time for DEKnull-2 subgroups to lose antibody response was estimated as 168 months. In addition, antibody clearance rates (adjusted per 100 persons-month) were similar between subgroups of DBPII-Sal1 (0.48 to 0.57) and DEKnull-2 (0.27 to 0.46) (S1Table).

**Fig 3.** Maintenance of *P. vivax* antibody response over time according to episodes of EBV-DNAemia (none, one or multiple). The Kaplan-Meier’s curves show the cumulative survival probabilities of maintaining an antibody response against (A) DBPII-Sal1 and (B) DEKnull-2 among responders who were stratified according to the presence or absence of detectable EBV-DNAemia. Each line represents the survival probability of antibodies over time among (i) individuals who did not have detectable EBV DNA episodes during the 14-years follow-up (dashed line); (ii) individuals who had a single EBV DNA episode (lilac line); and (iii) individuals who had more than one (multiple) EBV DNA episodes (purple line). Number of individuals and contribution time (persons/month) of each group were described in S1 Table.

### EBV serostatus and long-term *P. vivax* antibody response

To evaluate the EBV serological status of the study population, the presence and levels of IgG antibodies to EBV antigens associated with lytic (VCAp18) or latent (EBNA1) phases of infection were investigated. As expected for a predominantly adult population, responders’ frequencies were high (> 60%) for both viral proteins and it remained relatively stable until the end of the follow-up (S3 Fig). In general, IgG-VCAp18 levels were higher compared to IgG-EBNA1.

Spearman correlation analyses were performed to investigate whether the levels of EBV antibodies correlated with *P. vivax-*specific antibodies. Regardless of the malaria transmission period (high, I/III or low, II/IV), the results demonstrated that anti-EBV antibodies (VCAp18 and EBNA1) did not correlate with anti-*P. vivax* antibodies (DBPII-Sal1 and DEKnull-2) (Fig 4). Despite of that, the antibody responses to DBPII-Sal1 and DEKnull-2 were strongly correlated (r=0.71 to 0.82; p<0,0001 for all periods). In general, antibodies to VCAp18 and EBNA1 presented a weak but positive correlation (except for period IV).

**Fig 4.** Correlation between IgG antibody levels to *Plasmodium vivax* and EBV virus over time. Clustering was based on the Spearman correlation coefficient for assays measuring specific EBV antibodies (VCAp18 and EBNA1) and specific *P. vivax* antibodies (DBPII-Sal1 and DEKnull-2). Matrix heatmaps were illustrated for each study period (I, II, III and IV). Positive correlations are shown in blue and negative correlations are shown in red. The asterisks represent statistically significant differences (*p < 0,05; **p < 0,01; *** p < 0,001).

To determine whether EBV serological status could influence the long-term antibody response to DBPII-based antigens, IgG-VCAp18 and IgG-EBNA1 carriers were stratified as persistent (PR), temporary (TR) or non-responder (NR). Survival curves of antibodies allowed to demonstrate that the decline over time of responders to both DBPII-based antigens were similar between PR, TR or NR for either VCAp18 (Figs 5A and 5B) or EBNA1 (Figs 5C and 5D). Despite that, antibody clearance rates for DEKnull-2 responders (0.26 to 0.36 per 100 persons-month) declined more slowly than from DBPII-Sal1 (0.45 to 0.53 per 100 persons-months) (S1 Table).

**Fig 5.** Maintenance of *P. vivax* antibody response over time according to EBV serostatus (persistent-PR, temporary-TR, and non-responder-NR). The Kaplan-Meier’s curves show the cumulative survival probabilities of maintaining an antibody response against (A, C) DBPII-Sal1 and (B, D) DEKnull-2 among responders who were stratified according to the response profile for VCAp18 or EBNA1. Each line represents the survival probability of antibodies over time among (i) persistent responders, who had IgG response over the 14 years of follow-up (A and B - brown line; C and D - red line); (ii) temporary responders, who had variable IgG response during cross-sectional surveys (A and B - yellow line; C and D - pink line); and (iii) non-responders, who had no IgG response detected any time in the study (dashed line). Number of individuals and contribution time (person/month) of each group are described in S1 Table.

## Discussion

This 14-year follow-up study provided for the first time an exploratory description of the long-term *P. vivax* humoral immune response in the context of EBV co-infection. For this, immunogens based on the DBPII were used, including a common DBPII variant (Sal1) circulating in the Amazon area [16] and the engineered DEKnull-2 that has been associated with stronger, broader, and long-term neutralizing antibody response in the study area [18, 21].

In the study population, it was essential to identify viral DNA carriers, as circulating EBV DNA could be an indicator of active EBV replication [27]. In this P. *vivax* semi-immune adults, the detection of EBV DNA in peripheral blood decreased significantly over the years, precluding any attempts to classify individuals as long-term DNAemia carriers. The detection of viral DNA of most carriers did not persist over the first few months of the study, which may reflect that once infected by EBV early on, the subjects - in general - have good immunological control over EBV (reflected here by stable antibody responses to VCAp18 and EBNA1). Concomitantly, acute malaria infection (presence of parasites and/or *P. vivax* DNA in the peripheral blood) also decreased over time, which led to a positive association between the presence of acute *P. vivax* infection and EBV-DNAemia. At this point, it is not possible to determine whether this potential association represents a spurious correlation or may reflect an influence of concurrent *P. vivax* parasite in the activation of EBV replication cycle, as it has been proposed in the case of the more lethal malaria parasite, *P. falciparum* [28]. In Indonesia, where *P. falciparum* and *P. vivax* species are co-endemic at substantial proportions, it was demonstrated that EBV DNA levels were significantly elevated in both *P. falciparum* and *P. vivax* mono-infections as compared with uninfected controls, suggesting cytokine-mediated imbalance of immune control of EBV during active malaria [29]. Although the study from Indonesia supports our findings, the influence of concomitant transmission of *P. falciparum* cannot be ruled out in that area, especially because PCR-based protocols were not used to investigate submicroscopic infections. In the case of *P. falciparum* – whose relationship with EBV infections is involved in the genesis of endemic Burkitt lymphoma (eBL) [30] – it was demonstrated that repeated malaria infections were associated with increased proliferation and transformation of EBV-infected cells [31]. Also, the clearance of circulating EBV after *P. falciparum* antimalarial treatment suggests a direct relationship between active malaria infection and viral reactivation [28]. Of note, however, the aggressive eBL childhood cancer seems to be exclusively linked to *P. falciparum* exposure, but not to other human malaria parasites [32]. Because of this, the direct relationship between active *P. vivax* infection and viral reactivation needs further confirmation.

We further analyzed whether one or more episodes of viral DNAemia, at any time of the long-term follow-up period, could impact the persistence of *P. vivax*-specific antibody responses. Regardless of viral DNA carriers’ status, maintenance over time of antibody responses to either DBPII-Sal1 or DEKnull-2 was similar between individuals who had no, one or multiple episodes of EBV-DNAemia; for example, the average time for the loss of antibody response was 168 months for DEKnull-2 and from 156 to 168 months for DBPII-Sal1. In our previous case-control study, lower IgG reactivity to *P. vivax* antigens was detected in the group of persistent EBV DNA carriers (cases) compared to age-matched controls (no circulant viral DNA) [15]. Unfortunately, differences in the study design and the absence of a subset of persistent viral DNA carriers in the current study preclude a direct comparison with the previous result. Despite this, it must be noted that, in the present study, the frequencies of both malaria infection and circulating viral DNA progressively decreased over time, making it reasonable to speculate that this may have masked a possible influence of active EBV replication on the long-term *P. vivax* immune response.To properly address this topic, it would be of relevance to examine *P. vivax*-exposed children, as typically the EBV primary infection occurs during childhood, with an early EBV seroconversion in Latin American populations associated with low socioeconomic status [33].

To address whether EBV serostatus could influence acquired *P. vivax* antibody responses, individuals were further categorized as persistent (PR), temporary (TR), or non-responder (NR) for lytic (VCAp18) or latent (EBNA1) viral antigens. Although antibody clearance rates were faster for Sal-1 (0.49 to 0.55/100 persons-month) than for DEKnull-2 (0.27 to 0.37/100 persons-month), the results of the 14-year follow-up study demonstrated that probabilities of maintaining antibodies to DBPII-based antigens were similar for IgG-VCAp18 or IgG-EBNA1 subgroups (PR, TR, or NR). It is noteworthy that no association (positive or negative) was detected between the levels of anti-EBV antibodies and anti-*P. vivax* antibodies. Together, these results suggested that sustained EBV antibody responses to lytic/latent viral antigens did not impair the long-term antibody response to DBPII-related antigens, including both strain-specific (Sal1) and strain-transcending (DEKnull-2) immune responses.

Our study has limitations that should be considered when interpreting the results. First, a progressive decrease in malaria transmission over time precluded more robust statistical analysis to confirm the association between acute *P. vivax* infection and EBV-DNAemia. To explore whether concurrent *P. vivax* infection may result in EBV lytic replication, perhaps it should be more appropriate to evaluate the levels of viral DNA methylation, as unmethylated viral DNA occurs when EBV switches to the lytic phase and in virions [34]; nevertheless, the investigation of this topic was outside of the scope of the current study. Secondly, the EBV serostatus of the study population was constrained to the profile of IgG antibodies to VCAp18 (lytic) and EBNA1 (latent) antigens. Due to the limitations of plasma samples available over the 14 years of study, we chose the lytic antigen VCAp18, because from a panel of synthetic peptides targeting the most frequent EBV lytic antigens (i.e., IgG/IgM to VCAp18, IgM to ZEBRA and IgM EAd-p45/52), only the levels of anti-IgG VCAp18 showed a positive association with EBV DNA copies in the peripheral blood of Amazonian adults [15]. Notwithstanding these limitations, we are confident that, in this immunocompetent adult population, the assessment of VCAp18-IgG and EBNA1-IgG represented the best combination for long-term sero-detection of anti-EBV antibodies. Finally, as EBV latency establishment is in B cells [35], it should be of interest to investigate whether *P. vivax*-specific memory B cells are compromised during EBV reactivation.

In conclusion, in this 14-year follow-up study we demonstrate that, in an immunocompetent *P. vivax*-exposed adult population, neither sporadic episodes of circulating EBV DNA nor a sustained antibody response to lytic/latent EBV antigens influence the longevity of both strain-specific and strain-transcending DBPII immune responses. Future studies should investigate a possible association between the presence of acute *P. vivax* infection and EBV-DNAemia.

## Data Availability

All relevant data are within the manuscript and its Supporting Information files.

## Acknowledgments

We thank the inhabitants of Rio Pardo for enthusiastic participation in the study; the local malaria control team in Presidente Figueiredo for their logistic support; the Instituto Leônidas &. Maria Deane – Fiocruz-Amazônia and Fundação de Medicina Tropical Dr. Heitor Vieira Dourado for local support, and the Instituto René Rachou – Fiocruz-Minas, Belo Horizonte, MG, for overall support. LHC, FSK, and CFJF are research fellows from CNPq. Scholarship was sponsored by FAPEMIG (BAR), CAPES (LFFG and SLA) and the Fiocruz Innovation Support Initiative/Inova-Fiocruz (MFAN). We also thank the financial support from the Program for Institutional Internationalization of the Higher Education Institutions and Research Institutions of Brazil-CAPES-PrInt from FIOCRUZ. The authors thank the Network Technological Platforms from FIOCRUZ, for the support and financing of the services provided by Digital and Real-Time PCR Facility from René Rachou Institute - FIOCRUZ MG - RPT09D.

## Supporting information

**S1 Fig. Malaria infections in 77 eligible participants screened for the presence of circulanting EBV DNA during the follow-up study.** *P. vivax* infections are represented in red, *P. falciparum* in yellow, mixed infections (*P. falciparum* plus *P. vivax*) in orange, and uninfected samples in white. During the cross-sectional surveys, the presence or absence of episodes of EBV-DNAemia are represented in purple and white circles, respectively.

**S2 Fig. Dynamics of naturally acquired antibody responses to *P. vivax* DBPII-based antigens during the follow-up study.** (A) Frequencies and (B) levels of antibodies against DBPII-Sal1 and DEKnull-2, as determined by conventional serological assays (ELISA). Results were expressed as reactivity index (RI), with RI >1.0 considered as an ELISA-positive response. In B, black lines inside the violin plot represent the median RI, with light green and blue lines representing the interquartile range.

**S3 Fig. Dynamic of antibody responses to EBV peptides during the 14 years follow-up study.** (A) Frequencies and (B) levels of antibodies against VCAp18 and EBNA1, as determined by conventional serological assays (ELISA). Results were expressed as the optical density at 450 nm (OD450), with OD >0.37 and >0.20 considered as ELISA-positive responses to VCAp18 and EBNA1, respectively. In B, black lines inside the violin plot represent the median RI, with yellow and pink lines representing the interquartile range.

**S1 Table. DBPII-related antibody responses over time based on the presence or absence of EBV-DNAemia.**

**S2 Table. DBPII-related antibody responses over time based on the antibody response profile against EBV peptides**

